# Seroprevalence, seroconversion, and seroreversion of infection-induced SARS-CoV-2 antibodies among a cohort of children and adolescents in Montreal, Canada

**DOI:** 10.1101/2022.10.28.22281660

**Authors:** Kate Zinszer, Katia Charland, Laura Pierce, Adrien Saucier, Britt McKinnon, Marie-Ève Hamelin, Islem Cheriet, Margot Barbosa Da Torre, Julie Carbonneau, Cat Tuong Nguyen, Gaston De Serres, Jesse Papenburg, Guy Boivin, Caroline Quach

## Abstract

**Importance:** Repeated serological testing for SARS-CoV-2 allows the monitoring of antibody dynamics in populations, including detecting infections that are missed by RT-PCR or antigen testing. Understanding the factors associated with seroconversion and seroreversion as well as the duration of infection-induced antibodies can also inform public health recommendations regarding disease prevention and mitigation efforts.

**Objective:** To use serological testing to assess the prevalence, seroconversion, and seroreversion of infection-induced SARS-CoV-2 antibodies in children and adolescents in Montreal, Canada.

**Design:** This analysis reports on three rounds of data collection from a prospective cohort study (Enfants et COVID-19: Étude de séroprévalence [EnCORE]). The study rounds occurred as follows: Round 1 October 2020-March 2021, Round 2 May to July 2021, and Round 3 November 2021 to January 2022. Most Round 3 samples were collected prior to the spread of the Omicron BA.1 variant in Quebec.

**Setting:** Population-based sample.

**Participants:** Children and adolescents aged 2 to 17 years in Montreal, Canada.

**Exposure:** Potential exposure to SARS-CoV-2.

**Main Outcomes and Measures:** Participants provided dried blood spots (DBS) for antibody detection and parents completed online questionnaires for sociodemographics and COVID-19 symptoms and testing history. The serostatus of participants was determined by enzyme-linked immunosorbent assays (ELISAs) using the receptor-binding domain (RBD) from the spike protein and the nucleocapsid protein (N) as antigens. We estimated seroprevalence for each round of data collection and by participant and household characteristics. Seroconversion rates were calculated as were the likelihoods of remaining seropositive at six months and one year.

**Results:** The study included DBS samples from 1 632, 936, and 723 participants in the first, second, and third rounds of data collection, respectively. The baseline seroprevalence was 5·8% (95% CI 4·8-7·1), which increased to 10·5% and 10·9% for the respective follow-ups (95% CI 8·6-12·7; 95% CI 8·8-13·5). The overall average crude rate of seroconversion over the study period was 12·7 per 100 person-years (95% CI 10·9-14·5). Adjusted hazard rates of seroconversion by child and household characteristics showed higher rates in children who were female, whose parent identified as a racial or ethnic minority, and in households with incomes less than 100K. The likelihood of remaining seropositive at six months was 67% (95% CI 59-76) and dropped to 19% (95% CI 11%-33%) at one year.

**Conclusions and Relevance:** The data reported here provide estimates of pre-Omicron seroprevalence, seroconversion rates and time to seroreversion in a population-based cohort of children and adolescents. Serological studies continue to provide valuable contributions for infection prevalence estimates and help us better understand the dynamics of antibody levels following infection. Continued study of seroconversion and seroreversion can inform public health recommendations such as COVID-19 vaccination and booster schedules.

**KEY POINTS:** *Question:* What was the rate of seroconversion and time to seroreversion for SARS-CoV-2 antibodies among children and adolescents in Montreal between October 2020 to January 2022?

*Findings:* The overall average crude rate of seroconversion was 12·7 per 100 person-years (95% CI 10·9-14·5). We observed higher rates of seroconversion in children who were female, whose parent identified as a racial or ethnic minority, and in households with incomes less than 100K. Among all children who seroconverted, 71% had not been previously diagnosed with COVID-19. Median time to seroreversion was 7·5 months.

*Meaning:* Even before the emergence of the Omicron variants, we observed a high rate of seroconversion for infection-induced SARS-CoV-2 antibodies along with widespread antibody waning by one year. Many children and adolescents seroconverted despite not receiving a prior COVID-19 diagnosis, indicating that RT-PCR and antigen testing continue to underestimate true disease prevalence.

## INTRODUCTION

With the emergence of highly transmissible SARS-CoV-2 variants and widespread use of at-home tests, the COVID-19 pandemic continues to challenge public health surveillance efforts and limit the precision in disease occurrence estimates. In the absence of reliable data on rates of infection, seroprevalence studies remain important in estimating the prevalence of infected individuals, particularly among young and healthy populations who are more likely to experience mild or asymptomatic cases although contributing to disease transmission.

Repeated serological testing allows the monitoring of antibody dynamics in populations, including seroconversion and seroreversion. Several studies have documented these dynamics after SARS-CoV-2 infection, with general agreement that seroconversion, the development of antibodies specific to the virus, typically occurs within two to three weeks after symptom onset^1,2^ and children appear to be less likely to seroconvert than adults^3,4^. Among seropositive individuals, estimates of antibody waning are variable with some suggesting that seroreversion occurs more rapidly among younger adults and those with mild or asymptomatic infections.^5–8^ Few data are available on seroreversion in children, although a few cohort studies have found that IgG antibodies waned (yet remained detectable), starting at two months up to 18 months post-infection, with higher antibody levels consistently observed among younger children.^7,9–13^

In this prospective population-based cohort study, we used serological testing to assess the prevalence, seroconversion, and seroreversion of anti-SARS-CoV-2 antibodies in children and adolescents in Montreal, Canada. We also identified characteristics of study participants associated with increased risk of SARS-CoV-2 seropositivity.

## MATERIAL AND METHODS

### Study design and participants

Participants were initially recruited through selected day cares and schools in four different neighbourhoods. The neighborhoods were selected to reflect diversity in terms of geography, cumulative COVID-19 cases, and neighborhood socioeconomic status. The populations living in the West Island and Plateau-Mont-Royal are more affluent and highly educated, while Montreal North is one of the city’s poorest and most racially and ethnically diverse neighborhoods, and Mercier-Hochelaga-Maisonneuve is a working-class neighborhood with nearly one-third of the population living below the poverty line.^14^ Participants were eligible for enrollment if they attended one of the participating schools or day cares and if they were between the ages of 2 and 17 years. Multiple children per household were eligible if they all attended a participating school or day care.

We obtained electronic consent from parents or legal guardians and assent from children and teenage participants. The study protocol was approved by the research ethics boards of the Université de Montréal and the Centre Hospitalier Universitaire Sainte-Justine. Details of the full cohort study procedures are available in the study protocol.^15^ This study followed the reporting requirements of the Strengthening the Reporting of Observational Studies in Epidemiology (STROBE) statement.^16^

### Procedures

For each round of data collection, parents completed a questionnaire which included questions on the household and home, the general health of their child, any SARS-CoV-2 tests taken, observed symptoms, test type, and test results. Questions on household income and dwelling type were included after the baseline collection began. Once the questionnaires were complete, participants were sent specimen collection kits for finger prick whole blood samples. The present analysis includes data from three rounds of data collection: October 20, 2020, to March 15, 2021 (Round 1); May 18 to July 31, 2021 (Round 2); and November 18, 2021, to January 20, 2022 (Round 3).

### Laboratory testing

On reception in the laboratory, the samples were analyzed for quality control purposes, such as sufficient quantity of blood or layering of blood drops. For the samples that did not pass the quality control, participants and their parents were asked to redo the sample collection. The filter papers with dried blood spots (DBS) were conserved at −20 °C, and blood was eluted overnight the day before the serological assay. Samples were processed in 96-well plates with 40 samples from participants and 7 control samples (positive and negative) per plate.

The serostatus of participants was determined by a pair of enzyme-linked immunosorbent assays (ELISAs) that we developed and validated using the receptor-binding domain (RBD) from the spike protein and the nucleocapsid protein (N) as antigens. The ELISA assays were validated with positive control samples (participants with RT-PCR–confirmed SARS-CoV-2 infection and known to be seropositive for anti–SARS-CoV-2 antibodies) and negative control samples (SARS-CoV-2 seronegative). Based on the results, the RBD assay had a sensitivity of 95% and specificity of 100%, while the N assay had sensitivity of 90% and specificity of 95%. A colorimetric reaction determined by optic density (OD) allowed the detection of IgG in the samples and the evaluation of the signal generated by SARS-CoV-2–specific antibodies against the RBD and N antigens identified subjects as seropositive. The OD cutoff for positivity for each assay was determined based on the average of OD from negative sera plus 3 SDs. Samples collected in Round 1, prior to children’s vaccinations, were only tested using the RBD assay. Three samples, all from Round 1, had an indeterminate result (defined as a good quality sample near the OD cutoff) and were classified as seronegative. There were 59 seronegative samples that were initially identified as poor quality samples and subsequently analyzed as resubmitted samples were not received for these participants.

### Statistical analysis

Our original sample size calculation was based on seroprevalence estimates and we required a sample size of 457 children per neighborhood (total of 1 828 children) with a projected seroprevalence of 5%.^17^ Age was grouped to correspond to school level and vaccination age categories, i.e., 2-4, 5-11 and 12-17 years. Household income, a variable with several categories, was dichotomized as the lowest tercile vs the upper two terciles. The overweight binary variable was based on the CDC BMI percentile cut-off of 85%.^18^ For rates, confidence intervals were estimated based on the normal approximation to the Poisson distribution.^19^ All analyses were conducted using R version 4.2.0.

#### Seroprevalence estimates

If a participant was positive for RBD, was unvaccinated, or received their first dose of a COVID-19 vaccine within 9 days of their DBS sample, they were classified as being infection-induced seropositive.^20^ Participants who were both RBD and N positive were also classified as being infection-acquired seropositive, regardless of vaccination status. Those who were RBD positive but N negative, at least 9 days from the date of their first vaccination, were classified as vaccine-induced seropositive but seronegative for SARS-CoV-2 infection. Participants negative for both RBD and N, regardless of vaccination status, were classified as seronegative for SARS-CoV-2 infection. Seroprevalence was defined as the proportion of children that were seropositive following a SARS-CoV-2 infection among children providing a DBS sample of adequate quality. We estimated seroprevalence for each round of data collection and by participant and household characteristics. Unadjusted seroprevalence estimates and 95% confidence intervals (95% CI) were estimated by univariate logistic regression with inverse probability of censoring weight correction (IPCW) to correct for potential selection bias from loss to follow up. Significance was assessed with likelihood ratio tests (F-tests).^21^ IPCW was implemented using the ipw package and the survey package in R.^22^

#### Seroconversion estimates

Seroconversion rates were defined as the number of newly seropositive participants divided by the sum of participants’ time under observation while at-risk of infection, i.e., time to first seropositive test or time to first self-reported RT-PCR or antigen positive test, or time to last seronegative test for participants that remained uninfected for the duration of the follow-up period. Seroconversion rates were estimated for the full study period and separately for Rounds 2 and 3. The adjusted hazard rates of seroconversion were estimated using pooled logistic regression, an approach that is appropriate for non-proportional hazards and truncation.^23^ Missing values were imputed using Multivariate Imputation by Chained Equations (MICE).^24^ The minimally sufficient adjustment set of risk factors was identified using directed acyclic graphs constructed in DAGitty Version 3.0 (SFigures 1 and 2).^25^ The Variance Inflation Factor (VIF) was used to detect multicollinearity and any outliers were identified using Cook’s distance. When children were found to be seropositive at baseline, in the absence of a self-reported positive RT-PCR or antigen test, the approximate time of seroconversion was less accurately measured than for subsequent rounds of data collection. For this reason, a sensitivity analysis was conducted where the sample included only children seronegative at baseline or seropositive with a self-reported positive RT-PCR or antigen date. A separate sensitivity analysis was carried out using records with complete data (complete case analysis).

#### Seroreversion estimates

Seroreversion was defined as a seronegative test in a child that was previously seropositive. Days to seroreversion was defined as the number of days between the first positive test and the first negative serology test. If a positive self-reported RT-PCR test or antigen test date was provided and occurred prior to the seropositive test date, this self-report date was then used. The median time to seroreversion and the likelihood of remaining seropositive at six months and one year were estimated using Kaplan-Meier curves. To identify determinants of seroreversion we estimated adjusted hazard rates of seroreversion by pooled logistic regression.^23^ MICE was used to impute missing covariate data. Minimally sufficient adjustment sets, multicollinearity, and outliers were assessed as described earlier. In the main analyses, children seropositive in any round of data collection were followed to their first seronegative test or their last seropositive test if they never seroreverted. This required at least one serology test after seroconversion. Two sensitivity analyses were conducted: 1) excluding children seropositive at baseline without a self-reported RT-PCR or antigen positive test result and 2) censoring children vaccinated prior to first seronegative or last seropositive, at the time of vaccination. The first sensitivity analysis was motivated by the lack in accuracy of the time of seroconversion for seropositive children identified at the baseline assessment. For the second, the classification into seronegative status following vaccination is complex, therefore these results should provide a more accurate, though less statistically precise, estimate of time to seroreversion.

## RESULTS

There were 1 812 children, from 1 373 households, that initially enrolled in the study and completed the baseline questionnaire. Of these participants, 1 632 (90%) provided a DBS sample that was of sufficient quality for the serological analysis. Sixty-three additional children were recruited from 45 households and provided DBS in subsequent rounds. Fifty-seven percent of the participants were lost to follow-up by the third round of data collection but there was little evidence of differential loss to follow up by participant and household characteristics, with the exception of greater losses for participants from the West Island neighborhood and from households with parents who identified as a racial or ethnic minority (STable 1 in the supplement). The age group distribution changed through rounds of data collection mainly due to 2- to 4-year-olds aging into the 5- to 11-year group.

At baseline or Round 1 of data collection, the mean (SD) age of the children who provided a DBS sample was 9.6 (4.4) years, and 801 (49%) were female, with 354 participants (22%) from day cares, 725 (44%) from primary schools, and 553 (34%) from secondary schools. Most parents had at least a bachelor’s degree (1228 [75%]), and 201 (12%) self-identified as belonging to a racial or ethnic minority group (Table 1). On average, there were 2 adults and 2 children in 3 to 4–bedroom households. There were missing data values for household income for 10%-52% of children (depending on the round of data collection) and dwelling type 6%-46% of children. Bedroom density was missing for 23-25% of children, while BMI (height and/or weight) was missing or contained implausible values for 11-13% of children. The remaining variables had less than 2% missing values.

**Table 1.** Study population characteristics for each round of data collection.

|  | **Round 1**<br>**Children, No./**<br>**total No. (%)** | **Round 2**<br>**Children, No./**<br>**total No. (%)** | **Round 3**<br>**Children, No./**<br>**total No. (%)** |
| --- | --- | --- | --- |
| **Total** | 1632 | 936 | 723 |
| **Sex** |  |  |  |
| Female | 801/1632 (49.1) | 449/936 (49.0) | 342/723 (47.3) |
| Male | 831/1632 (50.9) | 487/936 (52.0) | 381/723 (52.7) |
| **Age, years** |  |  |  |
| 2-4 | 328/1632 (20.1) | 150/936 (16.0) | 88/723 (12.2) |
| 5-11 | 727/1632 (44.5) | 448/936 (47.9) | 346/723 (47.9) |
| 12-18 | 577/1632 (35.4) | 338/936 (36.1) | 289/723 (40.0) |
| **BMI** |  |  |  |
| Underweight or normal weight | 1179/1632 (72.2) | 687/936 (73.4) | 518/723 (71.6) |
| Overweight | 267/1632 (16.4) | 135/936 (14.4) | 109/723 (15.1) |
| **Chronic medical conditions** |  |  |  |
| None | 1517/1632 (93.0) | 861/936 (92.0) | 663/723 (91.7) |
| Yes | 103/1632 (6.3) | 69/936 (7.4) | 55/723 (7.6) |
| **Neighbourhood** |  |  |  |
| HOMA | 357/1632 (21.9) | 218/936 (23.3) | 173/723 (23.9) |
| Montreal North | 237/1632 (14.5) | 139/936 (14.9) | 121/723 (16.7) |
| Plateau | 534/1632 (32.7) | 302/936 (32.3) | 243/723 (33.6) |
| West Island | 504/1632 (30.9) | 277/936 (29.6) | 186/723 (25.7) |
| **Parental respondent's level of education** |  |  |  |
| No Bachelor's degree | 384/1632 (23.5) | 184/936 (19.7) | 166/723 (23.0) |
| Bachelor's degree | 647/1632 (39.6) | 380/936 (40.6) | 265/723 (36.7) |
| Graduate degree | 581/1632 (35.6) | 362/936 (38.7) | 284/723 (39.3) |
| **Bedroom density** |  |  |  |
| <1.5 pp bedroom | 875/1632 (53.6) | 513/936 (54.8) | 386/723 (53.4) |
| 1.5+ pp bedroom | 344/1632 (21.1) | 206 /936 (22.0) | 163/723 (22.5) |
| **Parental respondent's race and ethnicity** |  |  |  |
| Racial or ethnic minority | 201/1632 (12.3) | 110/936 (11.8) | 76/723 (10.5) |
| White | 1406/1632 (86.2) | 815/936 (87.1) | 640/723 (88.5) |
| **Number of bedrooms** |  |  |  |
| 1-2 bedrooms | 332/1632 (20.3) | 185/936 (19.8) | 154/723 (21.3) |
| 3 bedrooms | 740/1632 (45.3) | 417/936 (44.6) | 337/723 (46.6) |
| 4 bedrooms | 445/1632 (27.3) | 267/936 (28.5) | 182/723 (25.2) |
| 5 bedrooms | 104/1632 (6.4) | 61/936 (6.5) | 45/723 (6.2) |
| **Dwelling type** |  |  |  |
| Single family | 451/1632 (27.6) | 452/936 (48.3) | 294/723 (40.7) |
| Other | 424/1632 (26.0) | 425/936 (45.4) | 304/723 (42.0) |
| **Annual household income** |  |  |  |
| 0-100,000 | 246/1632 (15.1) | 263/936 (28.1) | 189/723 (26.1) |
| 100,000-150,000 | 551/1632 (33.3) | 576/936 (61.6) | 380/723 (52.5) |
| **HH member occupation** |  |  |  |
| Not essential | 913/1632 (55.9) | 515/936 (55.0) | 390/723 (53.9) |
| Essential, not health | 441/1632 (27.0) | 256/936 (27.4) | 203/723 (28.1) |
| Essential, health | 267/1632 (16.4) | 152/936 (16.2) | 120/723 (16.6) |

### Seroprevalence estimates

Serology data were available for 1 632, 936, and 723 children participating in the first, second, and third rounds of data collection, respectively. The Round 1 (October 2020 to March 2021) seroprevalence was 5.8% (95/1632, 95% CI 4.8-7.1) (Table 2). In Rounds 2 and 3, the seroprevalence had nearly doubled to 10.5% and 10.9%, respectively (93/936, 95% CI 8.6-12.7; 78/723, 95% CI 8.8-13.5). Participants living in single-family homes had significantly lower seroprevalence across all three rounds. The likelihood of seroprevalence was also significantly associated with neighborhood, with participants from the West Island having the lowest seroprevalence and Montreal North having the highest seroprevalence across all rounds. Though not consistently significant across all three rounds of data collection, participants in the 5- to 11-year-old age group, those with parents identifying as a racial or ethnic minority and those in homes with higher bedroom density and lower household income, tended to have higher seroprevalence.

**Table 2.** **Unadjusted seroprevalence by demographic characteristics by round of data collection weighted by inverse probability of censoring weights**

|  | Round 1 |  | Round 2 |  | Round 3 |  |
| --- | --- | --- | --- | --- | --- | --- |
|  | Seropositive Children, No./ total No. | Weighted Seroprevalence % (95% CI) | Seropositive Children, No./ total No. | Weighted Seroprevalence % (95% CI) | Seropositive Children, No./ total No. | Weighted Seroprevalence % (95% CI) |
| **Total** | 95/1632 | 5.8 (4.8-7.1) | 93/936 | 10.5 (8.6-12.7) | 78/723 | 10.9 (8.8-13.5) |
| **Sex** |  |  |  |  |  |  |
| Male | 40/831 | 4.8 (3.6-6.5) | 41/487 | 8.9 (6.6-11.9) | 33/381 | 8.7 (6.2-12.0) |
| Female | 55/801 | 6.9 (5.3-8.8) | 52/449 | 12.1 (9.3-15.5) | 45/342 | 13.2 (10.0-17.3) |
| **Age, years<sup>R3</sup>** |  |  |  |  |  |  |
| 2-4 | 16/328 | 4.9 (3.0-7.8) | 13/150 | 8.4 (4.9-14.0) | 6/88 | 6.3 (2.8-13.5) |
| 5-11 | 41/727 | 5.6 (4.2-7.6) | 54/448 | 12.9 (10.0-16.5) | 47/346 | 13.8 (10.5-18.0) |
| 12-18 | 38/577 | 6.6 (4.8-8.9) | 26/338 | 8.3 (5.7-12.0) | 25/289 | 9.0 (6.1-13.0) |
| **BMI** |  |  |  |  |  |  |
| Underweight or normal weight | 72/1179 | 6.1 (4.9-7.6) | 65/687 | 10.1 (8.0-12.7) | 59/518 | 11.4 (8.9-14.5) |
| Overweight | 12/267 | 4.5 (2.6-7.7) | 17/135 | 11.8 (7.4-18.4) | 8/109 | 7.5 (3.8-14.5) |
| **Chronic medical conditions** |  |  |  |  |  |  |
| None | 90/1517 | 5.9 (4.8-7.2) | 84/861 | 10.3 (8.3-12.6) | 70/663 | 10.8 (8.6-13.5) |
| Yes | 5/103 | 4.9 (2.0-11.1) | 8/69 | 12.3 (6.3-22.8) | 8/55 | 13.8 (7.0-25.5) |
| **Neighborhood<sup>R1, R2, R3</sup>** |  |  |  |  |  |  |
| HOMA | 28/357 | 7.8 (5.5-11.1) | 25/218 | 12.2 (8.3-17.6) | 20/173 | 12.5 (8.2-18.7) |
| Montreal North | 22/237 | 9.3 (6.2-13.7) | 22/139 | 17.0 (11.4-24.6) | 21/121 | 18.1 (12.0-26.4) |
| Plateau | 28/534 | 5.2 (3.6-7.5) | 29/302 | 9.8 (6.9-13.8) | 31/243 | 13.6 (9.7-18.8) |
| West Island | 17/504 | 3.4 (2.1-5.4) | 17/277 | 6.4 (4.0-10.2) | 6/186 | 3.0 (1.3-6.5) |
| **Parental respondent's level of education** |  |  |  |  |  |  |
| No Bachelor's degree | 18/384 | 4.7 (3.0-7.3) | 22/184 | 11.8 (7.8-17.5) | 16/166 | 8.6 (5.3-13.8) |
| Bachelor's degree | 41/647 | 6.3 (4.7-8.5) | 35/380 | 9.9 (7.2-13.5) | 28/265 | 10.7 (7.5-15.2) |
| Graduate degree | 36/581 | 6.2 (4.5-8.5) | 36/362 | 10.5 (7.6-14.3) | 34/284 | 12.9 (9.3-17.6) |
| **Parental respondent's race and ethnicity<sup>R1, R2</sup>** |  |  |  |  |  |  |
| Racial or ethnic minority | 22/201 | 10.9 (7.3-16.1) | 19/110 | 18.9 (12.3-27.9) | 10/76 | 13.4 (7.3-23.4) |
| White | 73/1406 | 5.2 (4.1-6.5) | 74/815 | 9.4 (7.5-11.7) | 67/640 | 10.4 (8.3-13.1) |
| **Number of bedrooms** |  |  |  |  |  |  |
| 1-2 bedrooms | 18/332 | 5.4 (3.4-8.4) | 22/185 | 12.3 (8.2-18.1) | 25/154 | 17.5 (12.0-24.6) |
| 3 bedrooms | 47/740 | 6.4 (4.8-8.4) | 40/417 | 10.1 (7.5-13.6) | 35/337 | 10.3 (7.4-14.1) |
| 4 bedrooms | 26/445 | 5.8 (4.0-8.4) | 27/267 | 10.7 (7.4-15.4) | 14/182 | 7.9 (4.7-13.0) |
| 5 bedrooms | 4/104 | 3.8 (1.5-9.8) | 4/61 | 6.6 (2.5-16.4) | 4/45 | 8.6 (3.2-20.9) |
| **Bedroom density<sup>R3</sup>** |  |  |  |  |  |  |
| [0,1.5) | 50/875 | 5.7 (4.4-7.5) | 51/513 | 10.5 (8.0-13.6) | 31/386 | 8.3 (5.9-11.7) |
| 1.5+ | 27/344 | 7.8 (5.4-11.2) | 27/206 | 13.6 (9.4-19.2) | 26/163 | 16.8 (11.7-23.7) |

**Table 2 Unadjusted seroprevalence by demographic characteristics by round of data collection weighted by inverse probability of censoring weights (continued)**
|  | Round 1 |  | Round 2 |  | Round 3 |  |
| --- | --- | --- | --- | --- | --- | --- |
|  | Seropositive Children, No./ total No. | Weighted Seroprevalence % (95% CI) | Seropositive Children, No./ total No. | Weighted Seroprevalence % (95% CI) | Seropositive Children, No./ total No. | Weighted Seroprevalence % (95% CI) |
| **Dwelling type**<sup>R1, R2, R3</sup> |  |  |  |  |  |  |
| Single family | 27/451 | 6.0 (4.1-8.6) | 36/452 | 7.9 (5.8-10.9) | 23/294 | 7.3 (4.8-10.8) |
| Other | 44/424 | 10.4 (7.8-13.7) | 49/425 | 12.6 (9.6-16.4) | 40/304 | 14.5 (10.8-19.2) |
| **HH members occupation** |  |  |  |  |  |  |
| Not essential | 44/913 | 4.8 (3.6-6.4) | 56/515 | 11.3 (8.7-14.4) | 39/390 | 10.0 (7.3-13.4) |
| Essential, not health | 30/441 | 6.8 (4.8-9.6) | 24/256 | 9.8 (6.6-14.3) | 24/203 | 12.5 (8.5-18.0) |
| Not essential | 21/267 | 7.9 (5.2-11.8) | 12/152 | 9.1 (5.1-15.7) | 15/120 | 12.2 (7.4-19.5) |
| **Annual household income**<sup>R1, R2</sup> |  |  |  |  |  |  |
| < 100,000 | 33/246 | 13.4 (9.7-18.3) | 37/263 | 15.2 (11.1-20.4) | 23/189 | 13.6 (9.1-19.7) |
| ≥ 100,000 | 36/551 | 6.5 (4.7-8.9) | 46/576 | 8.2 (6.2-10.9) | 38/380 | 10.0 (7.3-13.5) |

<sup>R1</sup> significant relationship with seroprevalence at baseline (R1)
<sup>R2</sup> significant relationship with seroprevalence in the second round of data collection (R2)
<sup>R3</sup> significant relationship with seroprevalence in the third round of data collection (R3)

### Seroconversion estimates

The overall average crude rate of seroconversion over the study period was 12.7 per 100 person-years (194 episodes in 1 523 person-years, 95% CI 10.9-14.5). In Round 2, the rate was 15.7 episodes in 100 person-years (67 episodes in 426 person-years, 95% CI 12.0-19.5) and in Round 3, the rate was 11.2 episodes in 100 person-years (32 in 287 person-years, 95% CI 7.3-15.0). Of the 194 infection-induced seroconversions, 137 (71%) did not have a positive self-reported RT-PCR or antigen test prior to the DBS. Across all rounds of data collection, there were 86 occurrences where a participant self-reported a positive RT-PCR or antigen test with 27 (31%) having had a negative serology test at least 14 days later.

Adjusted hazard rates of seroconversion by child and household characteristics showed higher rates in children who were female, whose parent identified as a racial or ethnic minority, and in households with incomes less than 100K (Table 3). The West Island had lower seroconversion rates compared to the other three neighborhoods. The sensitivity analyses with data on children seronegative at Round 1 or seropositive with a positive RT-PCR or antigen date and followed through two subsequent rounds of serology testing, did not find elevated seroconversion rates in females though other results were consistent with the main analysis (STable 3). The complete case analyses based on minimally sufficient adjustment sets were similar to the main seroconversion analysis findings (STable 4). Tests for multicollinearity and outliers indicated low correlation and no outliers.

**Table 3.** Adjusted relative likelihood of seroconversion through follow up.

|  | Full adjusted<br>N=1 690* |  | Minimally sufficient adjustment<br>N=1 690 |  |
| --- | --- | --- | --- | --- |
|  | Estimate<br>HR (95% CI) | P value | Estimate<br>HR (95% CI) | P value |
| **Sex** |  |  |  |  |
| Male | 1 [Reference] |  | 1 [Reference] |  |
| Female | 1.42 (1.05-1.92) | 0.02 | 1.48 (1.10-1.99) | 0.01 |
| **Age, years** |  |  |  |  |
| 2-4 | 1 [Reference] |  | 1 [Reference] |  |
| 5-11 | 1.22 (0.79-1.88) | 0.37 | 1.20 (0.79-1.83) | 0.39 |
| 12-18 | 0.99 (0.62-1.59) | 0.98 | 1.05 (0.67-1.63) | 0.84 |
| **BMI** |  |  |  |  |
| Underweight or normal weight | 1 [Reference] |  | 1 [Reference] |  |
| Overweight | 0.72 (0.45-1.16) | 0.18 | 0.81 (0.51-1.28) | 0.36 |
| **Chronic medical conditions** |  |  |  |  |
| None | 1 [Reference] |  | 1 [Reference] |  |
| Yes | 1.18 (0.68-2.08) | 0.56 | 1.17 (0.67-2.03) | 0.58 |
| **Neighbourhood** |  |  |  |  |
| Plateau | 1 [Reference] |  | 1 [Reference] |  |
| Montreal North | 1.46 (0.87-2.45) | 0.15 | 1.44 (0.87-2.37) | 0.16 |
| HOMA | 1.22 (0.81-1.84) | 0.35 | 1.14 (0.76-1.72) | 0.53 |
| West Island | 0.54 (0.30-0.95) | 0.03 | 0.51 (0.29-0.89) | 0.02 |
| **Parental respondent's level of education** |  |  |  |  |
| No Bachelor's degree | 1 [Reference] |  | 1 [Reference] |  |
| Bachelor's degree | 1.45 (0.93-2.26) | 0.10 | 1.09 (0.73-1.63) | 0.67 |
| Graduate degree | 1.59 (1.01-2.52) | 0.05 | 1.19 (0.80-1.77) | 0.40 |
| **Parental respondent's race and ethnicity** |  |  |  |  |
| White | 1 [Reference] |  | 1 [Reference] |  |
| Racial or ethnic minority | 1.60 (1.04-2.45) | 0.03 | 1.90 (1.29-2.79) | 0.001 |
| **Bedroom density (pp per bedroom)** |  |  |  |  |
| <1.5 | 1 [Reference] |  | 1 [Reference] |  |
| 1.5+ | 1.12 (0.79-1.61) | 0.52 | 1.27 (0.91-1.77) | 0.17 |
| **Dwelling type** |  |  |  |  |
| Other | 1 [Reference] |  |  |  |
| Single family | 1.09 (0.70-1.70) | 0.69 | 0.79 (0.57-1.11) | 0.17 |
| **Annual household income** |  |  |  |  |
| <100,000 | 1 [Reference] |  | 1 [Reference] |  |
| ≥100,000 | 0.61 (0.42-0.90) | 0.01 | 0.54 (0.38-0.77) | P<.001 |

**Table 3 Adjusted relative likelihood of seroconversion through follow up (continued)**
|  | Full adjusted<br>N=1 690* |  | Minimally sufficient adjustment<br>N=1 690 |  |
| --- | --- | --- | --- | --- |
|  | Estimate<br>HR (95% CI) | *P* value | Estimate<br>HR (95% CI) | *P* value |
| **HH member**<br>**occupation** |  |  |  |  |
| Not essential | 1 [Reference] |  | 1 [Reference] |  |
| Essential, not health | 1.08 (0.76-1.55) | 0.66 | 1.17 (0.83-1.65) | 0.37 |
| Essential | 1.16 (0.77-1.74) | 0.49 | 1.22 (0.82-1.82) | 0.33 |

\*longitudinal data from 1690 children; a minimum of 45 days of follow up (half the risk set period) was required for inclusion in the analysis

### Seroreversion estimates

In the main analysis, which included children who were seropositive during any round with at least one follow-up serology test, the median time to seroreversion was 228 days (95% CI 198-274)(Figure 1). There were 81 seroreversions, 41 in Round 2 and 40 in Round 3. The likelihood of remaining seropositive at six months (given that they had remained seropositive up to that point) was 67% (95% CI 59-76). The likelihood of remaining seropositive at one year dropped to 19% (95% CI 11%-33%). In the sensitivity analysis excluding seropositive children at baseline without a positive RT-PCR or antigen test date, the median time to seroreversion was 322 days and the likelihood of remaining positive at six months and one year was 70% and 30%, respectively. In the sensitivity analysis censoring vaccinated children at the time of vaccination, the median time to seroreversion was 249 days and the likelihood of remaining seropositive at six months and one year was 68% and 43%, respectively (STables 5 and 6). For the analyses of determinants of seroreversion, the number of participants contributing longitudinal data to the main pooled regression analysis was 145. In all adjusted main analyses and sensitivity analyses, there was no evidence of an association between the hazard rates and time to seroreversion and any of the factors under study, i.e., the presence of at least one symptom, having high body weight or obesity, having a chronic condition, round of data collection, neighbourhood, parent identification as an ethnic or racial minority, age, and sex (results not shown).

**Figure 1a.**
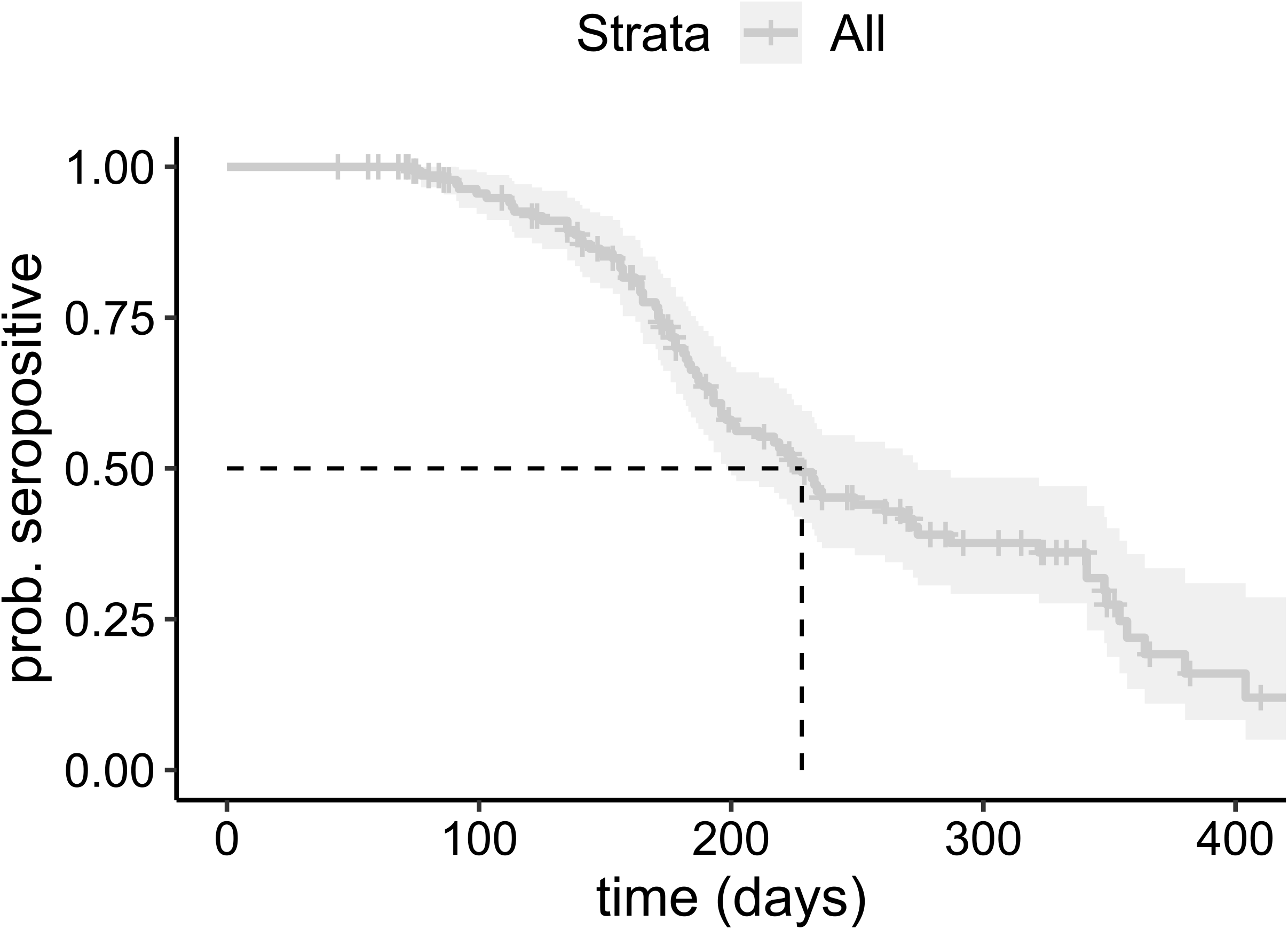
Kaplan-Meier curve of time to seroreversion using data from baseline and two follow-up assessments. The darker grey line represents the estimate with the light grey representing the 95% confidence intervals.

**Figure 1b.**
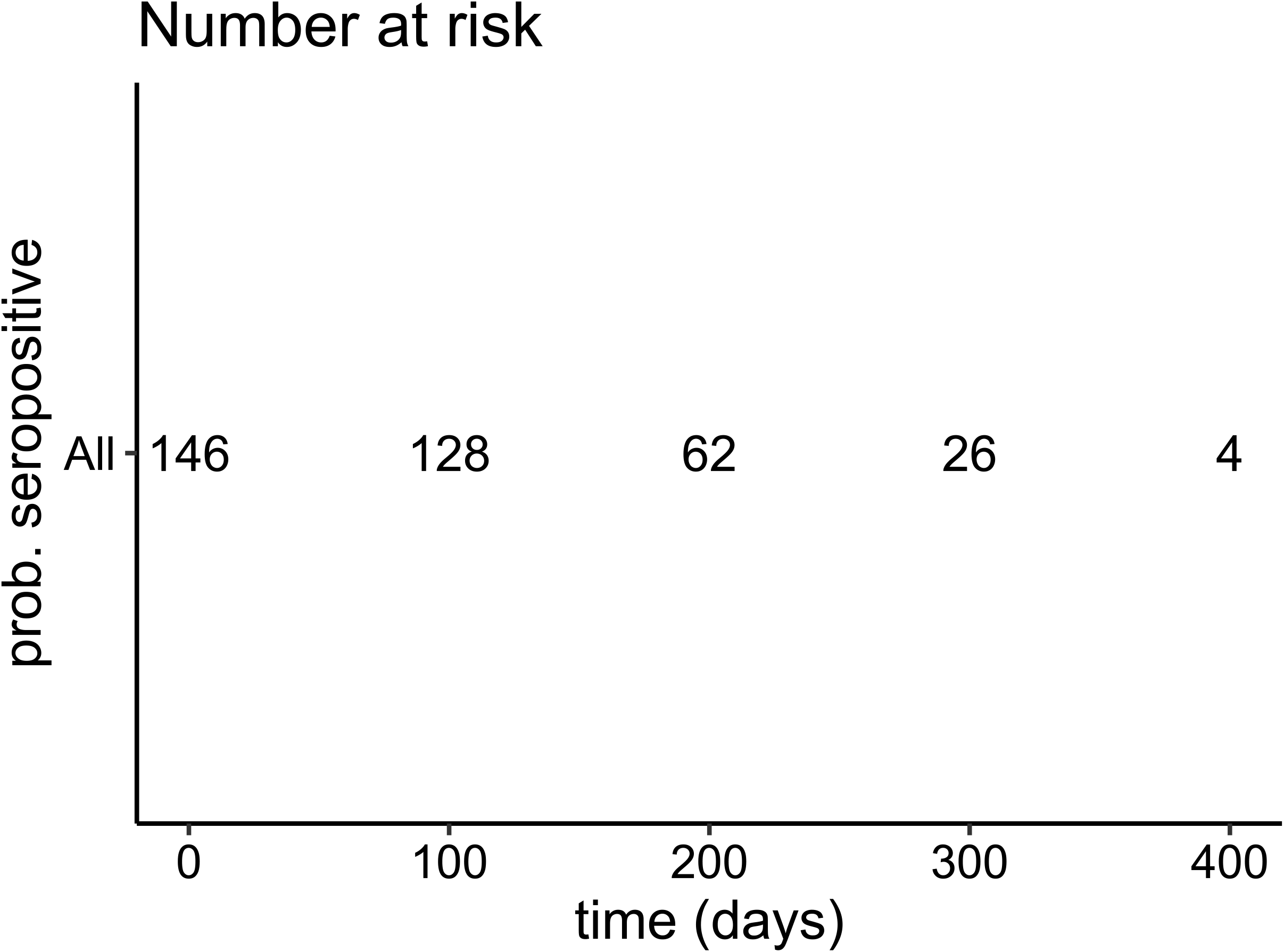
Number of participants at risk according to different time points. Figure represents number of seropositive participating children for 5 time points in days.

## DISCUSSION

The data reported here provide estimates of pre-Omicron seroprevalence, seroconversion rates and time to seroreversion, by household and participant characteristics, in a population-based cohort of children and adolescents from Montreal, Canada. We also identified significant risk factors for seroconversion over the study period. These results support other study findings in that the seroprevalence estimates increased over time and that the risk of seroconversion was significantly different in subgroups in regards to sex, household income, children of parents who identify as a racial or ethnic minority, household density, and neighbourhood of residence. It also provides further evidence of SARS-CoV-2 IgG antibody waning with a median seroreversion time of approximately 7.5 months and the likelihood of remaining seropositive at one year dropping to 19%.

The increase in seroprevalence over time was consistent with the pandemic context of Quebec as well as with other seroprevalence studies.^26,27^ A blood donor seroprevalence survey among Montreal adults had an estimated 6.4% seroprevalence of anti-N antibodies a few months prior to our Round 1 study period.^27^ For children from other regions of Canada, seroprevalence estimates from the pre-Omicron waves (waves 1-4) have been lower ranging from 2.8%^28^ to 4.6%^29^ while internationally, the estimates are higher in the United States with 44.2%^30^, England (20.9% to 36.3%)^26^, and 21.5%^10^ in Germany.

In our study, racial and ethnic minority status, higher household density, and lower household income was associated with increased risk of seropositivity and seroconversion in children. It has been established that there has been a disproportionate impact of the pandemic on racial and ethnic communities and lower income households due to social inequalities that have amplified the risk of infection including household crowding, type of occupation, and reduced access to healthcare.^31,32^ In our study population, female participants were at higher risk of seroconversion and seropositivity, which has not been found by most other studies.^33^ Participants from the West Island neighbourhood were consistently less at risk for seropositivity compared to the other neighbourhoods. In addition to the populations living in the West Island being more affluent and highly educated, compared to Montreal North and Mercier-Hochelaga-Maisonneuve, it is also a mainly residential suburb of the city, therefore having a lower population and household density. These disparities in social determinants of health likely attribute to the different seroprevalence estimates between the neighbourhoods.

In our study, 55% of our seropositive participants (that had at least two serology results) seroreverted. We also estimated that from the time of seroconversion, the likelihood of remaining seropositive at six months was 67% which dropped to 19% after one year. In other pediatric serological studies, the follow-up time has generally been shorter but studies have found that over time there is a waning of SARS-CoV-2 antibody levels.^11,34^ Other pediatric studies have not found an association with waning and asymptomic vs symptomatic infections^11^ but have with age^9^ and comorbidities such as respiratory disease and obesity. The waning appears to be more pronounced in adults and associated with obesity, older age, and belonging to a racial or ethnic minority.^35^

In this study, there was significant loss to follow-up and based on anecdotal reports from parents, this was largely associated with the DBS procedure. There was little difference in losses by household or participant characteristics. Correcting for selection bias in seroprevalence estimates by IPCW showed modest changes in seroprevalence point estimates providing support for a minor impact of loss to follow-up on our results.^36,37^ Our infection-acquired seroprevalence estimate among vaccinated children in Round 3 is likely underestimated due to lower sensitivity of the N assay compared with the RBD assay, and the possible more rapid waning of anti-N antibodies.^2,38–41^ Misclassification may also have occurred among unvaccinated children given the imperfect sensitivity and specificity of antibody testing. Though anti-RBD antibodies are relatively long-lasting, they appear more likely to fall below the threshold of detection than anti-S antibodies.^42^ Therefore, using only the RBD assay to determine seropositivity among unvaccinated children may have limited our ability to detect antibodies to infection in this group, although some studies suggest that RBD is more correlated with neutralizing antibodies and thus more correlated with protection.^43,44^

The study was powered for precision of neighborhood seroprevalence estimates, and given the substantial loss to follow-up, adjusted seroreversion and seroconversion analyses may have lacked power to identify important associations. The seroconversion analyses uncovered interesting associations with ethnic/racial minority, household income and neighborhood, but we did not find evidence of important associations with seroreversion. Serostatus could only be assessed periodically (e.g., on average at 6-month intervals), which resulted in error being introduced into the timing of seroconversion and seroreversion, and therefore, the adjusted hazard rates. As traditional time-to-event models were less appropriate to use, pooled logistic regression was well suited to this context of periodic assessments, by estimating then pooling hazard rates in successive intervals of the study period. Furthermore, there was likely measurement error in the estimation of the rates of seroconversion and the time to seroreversion, given that either event could have occurred weeks prior to or during the follow-up.

A major contribution of our study is the longitudinal nature of the study, which continues to be relevant given the introduction of variants and the waning of infection-induced antibodies. Given limited RT-PCR testing, serological studies provide valuable contributions for infection prevalence estimates in addition to helping us better understand the dynamics of antibody levels following infection, which can inform public health efforts and recommendations regarding risk mitigation and vaccination.^45^

## Data Availability

Individual participant data (after deidentification) and study documents will be available at study end. Researchers who wish to access the data should contact the corresponding author with a concise proposal for consideration. Researchers will be required to sign a data-access agreement.

## Figure headings and legends

**SFigure 1.**
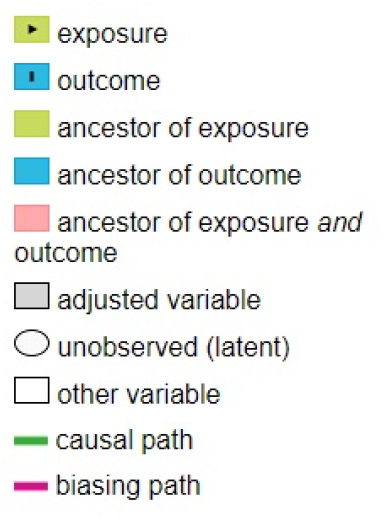
**Directed acyclic graph for seroconversion regression analysis to identify the minimally sufficient adjustment set for household income**

**SFigure 2.**
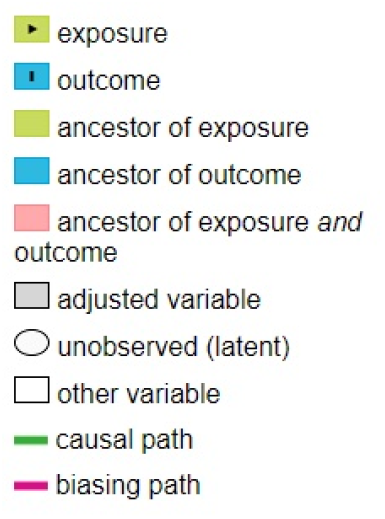
**Directed Acyclic Graph to determine the minimally sufficient adjustment set for the relationship of being symptomatic during infection and seroreversion**

## Author Contributions

Dr Zinszer had full access to all of the data in the study and takes responsibility for the integrity of the data and the accuracy of the data analysis.

*Concept and design:* Zinszer, Papenburg, De Serres, Quach.

*Acquisition, analysis, or interpretation of data:* Zinszer, Charland, Pierce, Saucier, McKinnon, Hamelin, Cheriet, Barbosa Da Torre, Carbonneau, Nguyen, De Serres, Boivin.

*Drafting of the manuscript*: Zinszer, Charland.

*Critical revision of the manuscript for important intellectual content:* Zinszer, Charland, Pierce, Saucier, McKinnon, Hamelin, Cheriet, Barbosa Da Torre, Carbonneau, Nguyen, De Serres, Papenburg, Boivin, Quach.

*Statistical analysis:* Charland, McKinnon.

*Obtained funding:* Zinszer, De Serres, Quach.

*Administrative, technical, or material support:* Zinszer, Charland, Pierce, Saucier, McKinnon, Hamelin, Cheriet, Barbosa Da Torre, Carbonneau, Nguyen, Boivin.

*Supervision*: Zinszer, Pierce, McKinnon, Boivin, De Serres.

## Roles of the Funder/Sponsor

The funder had no role in the design and conduct of the study; collection, management, analysis, and interpretation of the data; preparation, review, or approval of the manuscript; and decision to submit the manuscript for publication.

## Additional Contributions

We sincerely thank all children, teens, and their parents for their precious contribution to our study. We also thank the day cares, schools, and school boards for their help with recruitment. We are grateful for the support and guidance of our partners: the Direction régionale de santé publique de Montréal, the COVID-19 Immunity Task Force, the Institut national de santé publique du Québec, and the Observatoire pour l’éducation et la santé des enfants.

