## Supplemental Tables for "Seroprevalence, seroconversion, and seroreversion of infection-induced SARS-CoV-2 antibodies among a cohort of children and adolescents in Montreal, Canada"

**STable 1 Characteristics of children lost to follow up between first round and the third round of data collection**

|  | **Percentage lost to follow up** | **p-value** |
| --- | --- | --- |
| **Overall** | 57% |  |
| **Serostatus at baseline** |  |  |
| Positive | 46% | 0.02 |
| Negative | 58% |  |
| **Neighborhood** |  |  |
| HOMA | 54% | 0.01 |
| Montreal Nord | 55% |  |
| Plateau | 55% |  |
| West Island | 63% |  |
| **Racial or ethnic minority** |  |  |
| Yes | 63% | 0.04 |
| No | 56% |  |
| **Age category** |  |  |
| 2-4 | 59% | 0.64 |
| 5-11 | 57% |  |
| 12-18 | 58% |  |
| **Sex** |  |  |
| Male | 56% | 0.19 |
| Female | 59% |  |
| **Highest parent education** |  |  |
| less than Bachelors | 58% | 0.06 |
| Bachelors | 60% |  |
| Masters or higher | 53% |  |
| Missing | 62% |  |
| **Chronic condition** |  |  |
| Present | 49% | 0.19 |
| Absent | 58% |  |
| Missing | 62% |  |
| **Number of bedrooms** |  |  |
| 1-2 bedrooms | 55% | 0.42 |
| 3 bedrooms | 56% |  |
| 4 bedrooms | 61% |  |
| 5+ bedrooms | 57% |  |
| Missing | 58% |  |
| **Bedroom density** |  |  |
| [0,1.5) | 58% | 0.74 |
| 1.5+ | 56% |  |
| Missing | 58% |  |

**STable 1 Characteristics of children lost to follow up between first round and the third round of data collection (continued)**

|  | **Percentage lost to follow up** | **p-value** |
| --- | --- | --- |
| **Overweight** |  |  |
| Present | 61% | 0.09 |
| Absent | 58% |  |
| Missing | 51% |  |
| **Dwelling type** |  |  |
| Single family | 35% | NA |
| Other | 29% |  |
| Missing | 85% |  |
| **Household income** |  |  |
| <100K | 28% | NA |
| 100+K | 34% |  |
| Missing | 82% |  |
| **HH members occupation** |  |  |
| Not essential | 58% | 0.68 |
| Essential, not health | 57% |  |
| Essential, health | 56% |  |
| Missing | 47% |  |

*derived from questions asked in after baseline questionnaire began therefore p-values are not applicable

**STable 2 Unweighted, unadjusted seroprevalence by demographic characteristics by round of data collection for Round 2 and Round 3**

|  | **Round 2** | | **Round 3** | |
| --- | --- | --- | --- | --- |
|  | **Seropositive Children, No./ total No.** | **Weighted Seroprevalence**  **% (95% CI)** | **Seropositive Children, No./ total No.** | **Weighted Seroprevalence**  **% (95% CI)** |
| **Total** | 93/936 | 9.9 (8.2-12.0) | 78/723 | 10.8 (8.7-13.3) |
| **Sex** |  |  |  |  |
| Male | 41/487 | 8.4 (6.3-11.2) | 33/381 | 8.7 (6.2-11.9) |
| Female | 52/449 | 11.6 (8.9-14.9) | 45/342 | 13.2 (10.0-17.2) |
| **Age, years** |  |  |  |  |
| 2-4 | 13/150 | 8.7 (5.1-14.4) | 6/88 | 6.8 (3.1-14.4) |
| 5-11 | 54/448 | 12.1 (9.3-15.4) | 47/346 | 13.6 (10.4-17.6) |
| 12-18 | 26/338 | 7.7 (5.3-11.1) | 25/289 | 8.7 (5.9-12.5) |
| **BMI** |  |  |  |  |
| Underweight or normal weight | 65/687 | 9.5 (7.5-11.9) | 59/518 | 11.4 (8.9-14.4) |
| Overweight | 17/135 | 12.6 (8.0-19.3) | 8/109 | 7.3 (3.7-14.0) |
| **Chronic medical conditions** |  |  |  |  |
| None | 84/861 | 9.8 (7.9-11.9) | 70/663 | 10.6 (8.4-13.1) |
| Yes | 8/69 | 11.6 (5.9-21.5) | 8/55 | 14.5 (7.4-26.5) |
| **Neighbourhood** |  |  |  |  |
| HOMA | 25/218 | 11.5 (7.9-16.4) | 20/173 | 11.6 (7.6-17.2) |
| Montreal North | 22/139 | 15.8 (10.7-22.9) | 21/121 | 17.4 (11.6-25.2) |
| Plateau | 29/302 | 9.6 (6.8-13.5) | 31/243 | 12.8 (9.1-17.6) |
| West Island | 17/277 | 6.1 (3.8-9.6) | 6/186 | 3.2 (1.5-7.0) |
| **Parental respondent’s level of education** |  |  |  |  |
| No Bachelor’s degree | 22/184 | 12.0 (8.0-17.5) | 16/166 | 9.6 (6.0-15.2) |
| Bachelor’s degree | 35/380 | 9.2 (6.7-12.6) | 28/265 | 10.6 (7.4-14.9) |
| Graduate degree | 36/362 | 9.9 (7.3-13.5) | 34/284 | 12.0 (8.7-16.3) |
| **Parental respondent’s race and ethnicity** |  |  |  |  |
| Racial or ethnic minority | 19/110 | 17.3 (11.3-25.5) | 10/76 | 13.2 (7.2-22.8) |
| White | 74/815 | 9.1 (7.3-11.3) | 67/640 | 10.5 (8.3-13.1) |
| **Number of bedrooms** |  |  |  |  |
| 1-2 bedrooms | 22/185 | 11.9 (8.0-17.4) | 25/154 | 16.2 (11.2-22.9) |
| 3 bedrooms | 40/417 | 9.6 (7.1-12.8) | 35/337 | 10.4 (7.6-14.1) |
| 4 bedrooms | 27/267 | 10.1 (7.0-14.3) | 14/182 | 7.7 (4.6-12.6) |
| 5 bedrooms | 4/61 | 6.6 (2.5-16.2) | 4/45 | 8.9 (3.4-21.4) |
| **Bedroom density** |  |  |  |  |
| [0,1.5) | 51/513 | 9.9 (7.6-12.8) | 31/386 | 8.0 (5.7-11.2) |
| 1.5+ | 27/206 | 13.1 (9.1-18.4) | 26/163 | 16.0 (11.1-22.4) |

**STable 2 Unweighted, unadjusted seroprevalence by demographic characteristics by round of data collection for Round 2 and Round 3 (continued)**

|  | **Round 2** | | **Round 3** | |
| --- | --- | --- | --- | --- |
|  | **Seropositive Children, No./ total No.** | **Unweighted Seroprevalence**  **% (95% CI)** | **Seropositive Children, No./ total No.** | **Unweighted Seroprevalence**  **% (95% CI)** |
| **Dwelling type** |  |  |  |  |
| Single family | 36/452 | 8.0 (5.8-10.8) | 23/294 | 7.8 (5.3-11.5) |
| Other | 49/425 | 11.5 (8.8-14.9) | 40/304 | 13.2 (9.8-17.4) |
| **HH members occupation** |  |  |  |  |
| Not essential | 56/515 | 10.9 (8.5, 13.9) | 39/390 | 10.0 (7.4, 13.4) |
| Essential, not health | 24/256 | 9.4 (6.4, 13.6) | 24/203 | 11.8 (8.1, 17.0) |
| Essential, health | 12/152 | 7.9 (4.5, 13.4) | 15/120 | 12.5 (7.7, 19.7) |
| **Annual household income** |  |  |  |  |
| < 100,000 | 37/263 | 14.1 (10.4-18.8) | 23/189 | 12.2 (8.2-17.6) |
| ≥ 100,000 | 46/576 | 8.0 (6.0-10.5) | 38/380 | 10.0 (7.4-13.4) |

**STable 3 Sensitivity analysis of adjusted relative likelihood of seroconversion children PCR positive or seronegative at baseline followed over two follow-up rounds of data collection**

|  | **Fully adjusted**  **N=961*** | | **Minimally sufficient adjustment set**  **N=961** | |
| --- | --- | --- | --- | --- |
|  | **Estimate**  **HR (95% CI)** | ***P* value** | **Estimate**  **HR (95% CI)** | ***P* value** |
| **Sex** |  |  |  |  |
| Male | 1 [Reference] |  | 1 [Reference] |  |
| Female | 1.28 (0.86-1.90) | 0.22 | 1.35 (0.93-1.98) | 0.12 |
| **Age, years** |  |  |  |  |
| 2-4 | 1 [Reference] |  | 1 [Reference] |  |
| 5-11 | 1.42 (0.82-2.46) | 0.21 | 1.27 (0.75-2.15) | 0.37 |
| 12-18 | 0.82 (0.43-1.54) | 0.53 | 0.74 (0.41-1.33) | 0.31 |
| **BMI** |  |  |  |  |
| Underweight or normal weight | 1 [Reference] |  | 1 [Reference] |  |
| Overweight | 1.09 (0.64-1.87) | 0.74 | 1.22 (0.73-2.04) | 0.45 |
| **Chronic medical conditions** |  |  |  |  |
| None | 1 [Reference] |  | 1 [Reference] |  |
| Yes | 1.55 (0.81-2.94) | 0.18 | 1.50 (0.80-2.82) | 0.21 |
| **Neighbourhood** |  |  |  |  |
| Plateau | 1 [Reference] |  | 1 [Reference] |  |
| Montreal North | 1.38 (0.70-2.69) | 0.35 | 1.40 (0.73-2.66) | 0.31 |
| HOMA | 1.20 (0.71-2.04) | 0.50 | 1.17 (0.69, 1.97) | 0.56 |
| West Island | 0.37 (0.17-0.81) | 0.01 | 0.37 (0.17-0.79) | 0.01 |
| **Parental respondent’s level of education** |  |  |  |  |
| No Bachelor’s degree | 1 [Reference] |  | 1 [Reference] |  |
| Bachelor’s degree | 1.33 (0.74-2.38) | 0.34 | 0.99 (0.59-1.67) | 0.97 |
| Graduate degree | 1.51 (0.84-2.72) | 0.17 | 1.16 (0.69-1.93) | 0.58 |
| **Parental respondent’s race and ethnicity** |  |  |  |  |
| White | 1 [Reference] |  | 1 [Reference] |  |
| Racial or ethnic minority | 1.47 (0.83-2.62) | 0.19 | 1.81 (1.07-3.05) | 0.027 |
| **Bedroom density (pp per bedroom)** |  |  |  |  |
| <1.5 | 1 [Reference] |  | 1 [Reference] |  |
| 1.5+ | 1.15 (0.74, 1.81) | 0.53 | 1.43 (0.93-2.18) | 0.10 |
| **Dwelling type** |  |  |  |  |
| Other | 1 [Reference] |  |  |  |
| Single family | 1.10 (0.62-1.93) | 0.75 | 0.70 (0.45-1.08) | 0.11 |
| **Annual household income** |  |  |  |  |
| <100,000 | 1 [Reference] |  | 1 [Reference] |  |
| ≥100,000 | 0.69 (0.43-1.12) | 0.13 | 0.61 (0.39-0.95) | 0.03 |

**STable 3 Sensitivity analysis of adjusted relative likelihood of seroconversion children PCR positive or seronegative at baseline followed over two follow-up rounds of data collection (continued)**

|  | **Fully adjusted**  **N=961*** | | **Minimally sufficient adjustment set**  **N=961** | |
| --- | --- | --- | --- | --- |
|  | **Estimate**  **HR (95% CI)** | ***P* value** | **Estimate**  **HR (95% CI)** | ***P* value** |
| **HH member occupation** |  |  |  |  |
| Not essential | 1 [Reference] |  | 1 [Reference] |  |
| Essential, not health | 0.98 (0.61-1.57) | 0.94 | 1.02 (0.65-1.59) | 0.93 |
| Essential | 0.92 (0.53-1.58) | 0.76 | 1.02 (0.60-1.74) | 0.93 |

*longitudinal observations from 961 children

**STable 4 Sensitivity analysis of seroconversion hazard rates for participants with complete covariate data**

|  | **Full adjusted**  **N=530*** | | **Minimally sufficient adjustment set**  **N=530*** | |
| --- | --- | --- | --- | --- |
|  | **Estimate**  **HR (95% CI)** | ***P* value** | **Estimate**  **HR (95% CI)** | ***P* value** |
| **Sex** |  |  |  |  |
| Male | 1 [Reference] |  | 1 [Reference] |  |
| Female | 2.12 (1.38-3.31) | 0.001 | 1.48 (1.10-2.00) | 0.01 |
| **Age, years** |  |  |  |  |
| 2-4 | 1 [Reference] |  | 1 [Reference] |  |
| 5-11 | 1.15 (0.67-2.03) | 0.63 | 1.20 (0.80-1.85) | 0.39 |
| 12-18 | 0.85 (0.47-1.57) | 0.59 | 1.05 (0.68-1.65) | 0.84 |
| **BMI** |  |  |  |  |
| Underweight or normal weight | 1 [Reference] |  | 1 [Reference] |  |
| Overweight | 0.79 (0.40-1.44) | 0.47 | 0.93 (0.54-1.55) | 0.80 |
| **Chronic medical conditions** |  |  |  |  |
| None | 1 [Reference] |  | 1 [Reference] |  |
| Yes | 0.85 (0.29-2.03) | 0.74 | 0.80 (0.33-1.66) | 0.58 |
| **Neighbourhood** |  |  |  |  |
| Plateau | 1 [Reference] |  | 1 [Reference] |  |
| Montreal North | 1.33 (0.60-2.88) | 0.47 | 1.50 (0.79-2.79) | 0.21 |
| HOMA | 1.28 (0.75-2.17) | 0.37 | 1.14 (0.72-1.80) | 0.54 |
| West Island | 0.51 (0.24-1.06) | 0.07 | 0.42 (0.21-0.79) | 0.008 |
| **Parental respondent’s level of education** |  |  |  |  |
| No Bachelor’s degree | 1 [Reference] |  | 1 [Reference] |  |
| Bachelor’s degree | 2.20 (1.11-4.64) | 0.03 | 1.09 (0.74-1.64) | 0.66 |
| Graduate degree | 2.27 (1.15-4.82) | 0.02 | 1.18 (0.80-1.77) | 0.42 |
| **Parental respondent’s race and ethnicity** |  |  |  |  |
| White | 1 [Reference] |  | 1 [Reference] |  |
| Racial or ethnic minority | 1.76 (0.97-3.09) | 0.06 | 1.91 (1.28-2.77) | 0.001 |
| **Bedroom density (pp per bedroom)** |  |  |  |  |
| <1.5 | 1 [Reference] |  | 1 [Reference] |  |
| 1.5+ | 1.27 (0.80, 1.99) | 0.31 | 1.21 (0.80, 1.79) | 0.35 |
| **Dwelling type** |  |  |  |  |
| Other | 1 [Reference] |  | 1 [Reference] |  |
| Single family | 1.14 (0.67-1.92) | 0.62 | 0.77 (0.53-1.11) | 0.16 |
| **Annual household income** |  |  |  |  |
| <100,000 | 1 [Reference] |  | 1 [Reference] |  |
| ≥100,000 | 0.58 (0.36-0.94) | 0.03 | 0.54 (0.37-0.78) | 0.001 |
| **HH member occupation** |  |  |  |  |
| Not essential | 1 [Reference] |  | 1 [Reference] |  |
| Essential, not health | 1.10 (0.67-1.78) | 0.71 | 1.16 (0.82-1.64) | 0.38 |
| Essential | 0.92 (0.50-1.61) | 0.77 | 1.23 (0.81-1.82) | 0.31 |

*longitudinal observations from 530 children

**STable 5 Sensitivity analysis for median days to seroreversion with 95% confidence intervals***

| **Sample of seropositive children** | **n** | **events** | **Median days to seroreversion** | **Lower 95% CI** | **Upper 95% CI** |
| --- | --- | --- | --- | --- | --- |
| Seropositive at baseline or follow-up | 146 | 81 | 228 days | 198 days | 274 days |
| Seropositive at follow-up assessments or at baseline with self-reported positive test date | 84 | 31 | 322 days | 228 days | NA |
| Excluding children vaccinated before seroconversion and censoring children vaccinated after seroconversion at vaccination + 10 days | 141 | 53 | 233 days | 200 days | NA |

*All subjects in the analysis had seroconverted but time from seroconversion was based on self-reported PCR test, when available

**STable 6 Sensitivity analysis for likelihood of remaining seropositive at six month and one year with 95% confidence intervals ***

| **Sample of seropositive children** | **Elapsed Time (days)** | **Likelihood seropositivity** | **Lower 95% CI** | **Upper 95% CI** |
| --- | --- | --- | --- | --- |
| Seropositive at baseline or follow-up | 183 | 67% | 59% | 76% |
|  | 365 | 19% | 11% | 33% |
| Seropositive at follow-up assessments or at baseline with self-reported positive test date | 183 | 71% | 60% | 83% |
|  | 365 | 30% | 16% | 57% |
| Excluding children vaccinated before seroconversion and censoring children vaccinated after seroconversion at vaccination + 10 days | 183 | 68% | 59% | 77% |
|  | 365 | 41% | 31% | 55% |

*All subjects in the analysis had seroconverted but time from seroconversion was based on self-reported PCR test, when available
