## Supplemental Figure 1 for "Seroprevalence, seroconversion, and seroreversion of infection-induced SARS-CoV-2 antibodies among a cohort of children and adolescents in Montreal, Canada"

**SFigure 1 Directed acyclic graph for seroconversion regression analysis to identify the minimally sufficient adjustment set for household income**

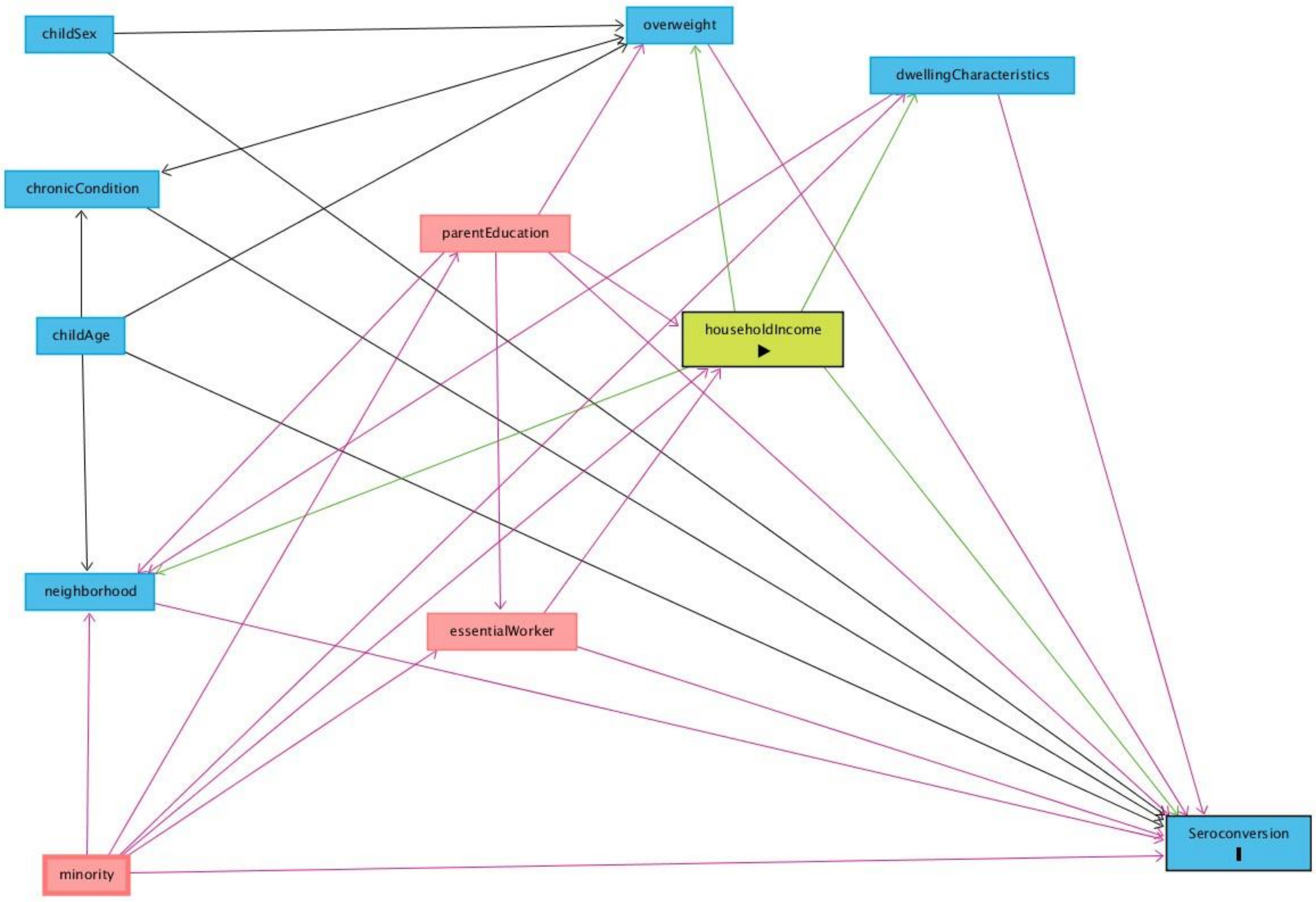
