## Supplemental Figure 2 for "Seroprevalence, seroconversion, and seroreversion of infection-induced SARS-CoV-2 antibodies among a cohort of children and adolescents in Montreal, Canada"

**SFigure 2 Directed Acyclic Graph to determine the minimally sufficient adjustment set for the relationship of being symptomatic during infection and seroreversion**

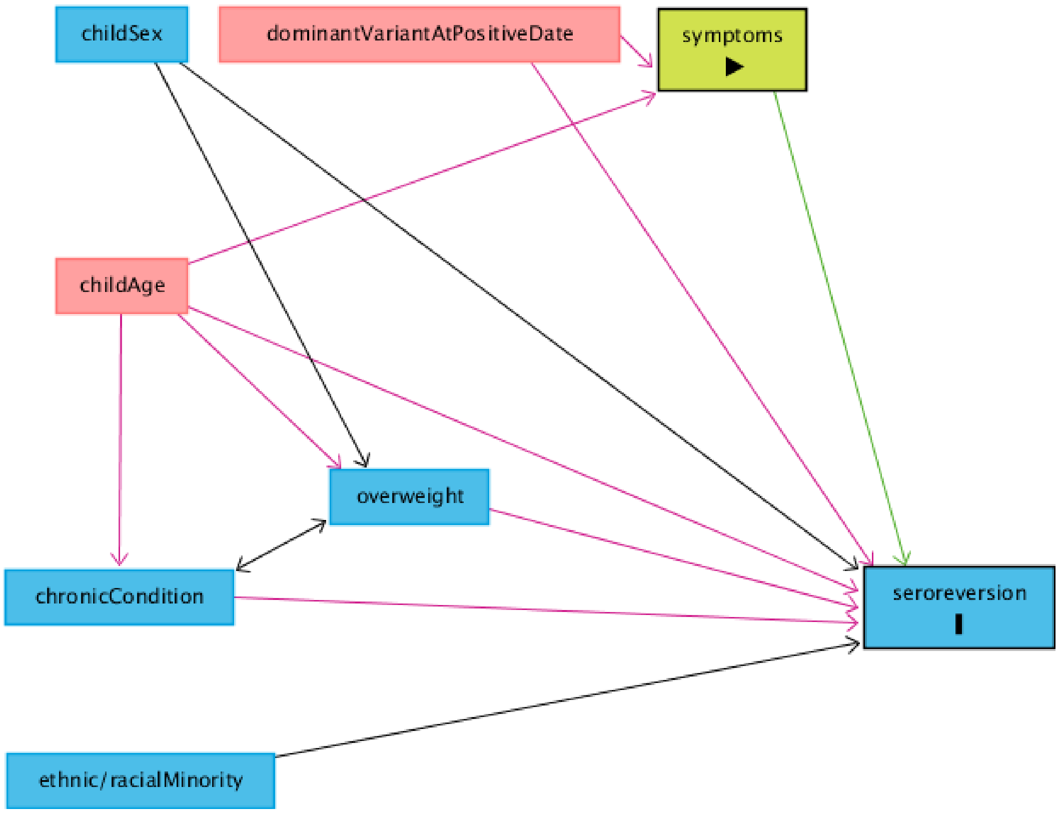
